# The effect of pre-booked appointments on COVID-19 vaccine uptake during the 2023 autumn campaign in the Netherlands: a regression discontinuity analysis

**DOI:** 10.1101/2025.07.10.25331000

**Authors:** Manon R. Haverkate, Jan van de Kassteele, Susan van den Hof, Jet G. Sanders, Mattijs S. Lambooij, Marijn de Bruin, Hester E. de Melker, Susan J.M. Hahné

**Affiliations:** Dutch National Institute for Public Health and the Environment (RIVM), Bilthoven, the Netherlands; Leiden University Medical Centre, Leiden, the Netherlands; London School of Economics and Political Science, London, United Kingdom; Erasmus School of Health Policy and Management, Erasmus University, Rotterdam, the Netherlands

## Abstract

**Introduction:** There is evidence that pre-booked appointments (PBA) for vaccination can enhance uptake, but might also result in reactance. We assessed the effect of PBA (versus self-scheduling) on uptake of COVID-19 vaccination during the 2023 autumn campaign in the Netherlands.

**Methods:** Persons aged ≥60 years were personally invited by mail. Those born between 01-01-1934 and 01-05-1952 (i.e., age 71.7-90.0 on 31-12-2023) received a letter with a PBA, while the others received a letter inviting them to self-schedule an appointment. National registries of sociodemographic determinants and COVID-19 vaccination were linked by a unique personal identifier. A regression discontinuity design was applied to estimate the local average treatment effect at 71.7 and 90.0 years of age. Stratified analyses were done for sociodemographic subgroups.

**Results:** The autumn 2023 vaccination coverage among persons aged ≥60 years with ≥1 prior registered COVID-19 vaccination (N = 4.0 million) was 55.0%. PBA resulted in a 3.3 (95% CI 2.6-4.1) and 4.5 (95% CI 2.8-6.3) percentage point higher uptake at 71.7 and 90.0 years, respectively. Subgroup analyses showed similar results. In one subgroup of migrants a negative effect of PBA was found, however, the confidence interval was wide.

**Discussion:** This nationwide quasi-experimental study shows that PBA is predominantly effective in increasing uptake by reducing the intention-behaviour gap. However, differences between subgroups should be taken into account to increase equity of the vaccination programme. If PBA leads to a lower uptake in certain subgroups with already a relatively low uptake, disparities likely become larger.

## Introduction

Within one year into the COVID-19 pandemic, vaccines became available to the public. In the Netherlands, vaccination started on January 6, 2021 [1]. All Dutch inhabitants aged 18 and over received a personal letter by mail, inviting them to self-schedule an appointment. Vaccine effectiveness of COVID-19 vaccination was high but decreased over time [2]. Consequently, several vaccination rounds have been organised afterwards. After the primary series and (multiple) booster vaccinations in 2021 and 2022, the first yearly autumn campaign was organised in the Netherlands in 2022 [3]. This vaccination campaign aimed to protect the Dutch population against the expected COVID-19 upsurge in the winter. In the 2022 autumn campaign, all Dutch inhabitants could receive a COVID-vaccination, although the main focus was on elderly and persons with underlying medical conditions, as they are at higher risk of severe disease, hospitalisation and death after SARS-CoV-2 infection, and vaccination is effective to prevent this [2, 4]. During the 2022 autumn campaign, persons aged 60 years and over and persons below the age of 60 with a medical condition received a letter inviting them to self-schedule an appointment, from the Dutch National Institute for Public Health and the Environment (RIVM) or their GP, respectively. All other Dutch inhabitants were informed through the media (e.g., television and radio commercials, social media advertisements) about when they could book their vaccination appointment.

In autumn 2023 another vaccination round was organised [5]. That year, vaccination was only available to those most at risk: persons aged 60 years and over on 31 December 2023; and those below 60 years of age with a medical condition. Only persons aged 60 years and over were personally invited by mail by the RIVM. They either received a letter with a pre-booked appointment (those born between 01-01-1934 and 01-05-1952), or a letter inviting them to self-schedule an appointment (all others aged 60 years and over). The aim of pre-booked appointments was changing the default: by presenting them with an appointment for COVID-19 vaccination, the hypothesis was that people would be more inclined to get vaccinated. They could still reschedule or cancel the appointment. This is deemed the most effective form of nudging: an intervention that makes the desired option more pronounced, without prohibiting the alternative [6].

There are many studies exploring the effects of nudging with the aim of increasing COVID-19 vaccine uptake. For example, text-based reminders [7], video-based messages [8, 9], granting freedoms [10], financial incentives [9–11], vaccination at local doctors [10], emphasising ease-of-access [9, 12], and presenting a social norm [13] have been used to nudge people toward vaccination. The nudge of sending letters with pre-booked appointments versus self-scheduling for COVID-19 vaccination was previously studied on a small scale in Sweden, Italy, and the US [14–17]. All studies found a predominantly positive effect of pre-booked appointments compared to self-scheduling an appointment on vaccine uptake. This nudge has also shown effective in other healthcare settings, for example in cancer screening programmes [18, 19].

One concern with respect to changing the default is reactance in a population [20, 21]. Whilst pre-scheduled appointments may make vaccinating for those who want to easier, for those in doubt it may feel like government interferes with their ‘free choice’ [20]. This is particularly sensitive for COVID-19 vaccination as it was a much more polarizing topic than most other choice contexts which have implemented default interventions. For this reason, prior to the implementation of this policy in the Netherlands in autumn of 2023, Sanders et al. [22] tested effects of different invitation systems on preferences and vaccination intention. They concluded that changing the default reduces the well-known gap between vaccination intention and behaviour [23], without affecting willingness to get vaccinated amongst the risk group.

Subsequently, we aim to assess the effect of pre-booked appointments on the uptake of COVID-19 vaccination during the 2023 autumn campaign in the Netherlands in persons aged 60 years and older using a regression discontinuity design (RDD). RDD is a quasi-experimental approach that allows for estimation of the causal effect of an exposure without the need of a randomized clinical trial [24–28]. Real-world data at population scale can be used to estimate the effect of an intervention or policy change. Furthermore, we will assess whether the effect of the invitation strategy is different for subgroups through stratification by, for example, previous COVID-19-vaccination, country of origin and socio-economic status. This can give valuable information on possible targeted invitation strategies, aiming to reduce inequity in the vaccination programme.

## Methods

### Study design

An observational cross-sectional study was performed using routinely collected COVID-19 vaccination registry data. The 2023 autumn vaccination campaign started on October 2, 2023 and ended on December 31, 2023.

### Population

The population consisted of the Dutch population aged 60 years and older on 31 December 2023, registered in the national Personal Records Database (N = 4,920,263; Figure 1). Persons born between 01-01-1934 and 01-05-1952 – i.e., age 71.7 to 90.0 on 31-12-2023 – could receive a letter with a pre-booked appointment (PBA) when they fulfilled the below eligibility criteria, while all others received a letter inviting them to self-schedule an appointment. For the exact letter content and how the letters were developed see Sanders et al. [22]. Persons were eligible for a PBA letter if they were previously vaccinated by the municipal health services (MHS) at least once, were registered in the COVID-vaccination Information and Monitoring System (CIMS), and had their last vaccination at least 3 months before the start date of the campaign. Thus, all persons who were previously vaccinated, but only by other providers (e.g., general practitioner, elderly care physician), those who were not vaccinated or did not give consent to be registered in CIMS, and those vaccinated within 3 months before the start of the campaign, received a letter asking them to self-schedule an appointment and were excluded from the analyses. Also, if no appointment could be made for a person eligible for a PBA letter (i.e., there was no PBA location available within 30 kilometres or there was no time slot available at a nearby location), a letter asking them to self-schedule an appointment was sent and these persons were also excluded from the analyses.

**Figure 1.**
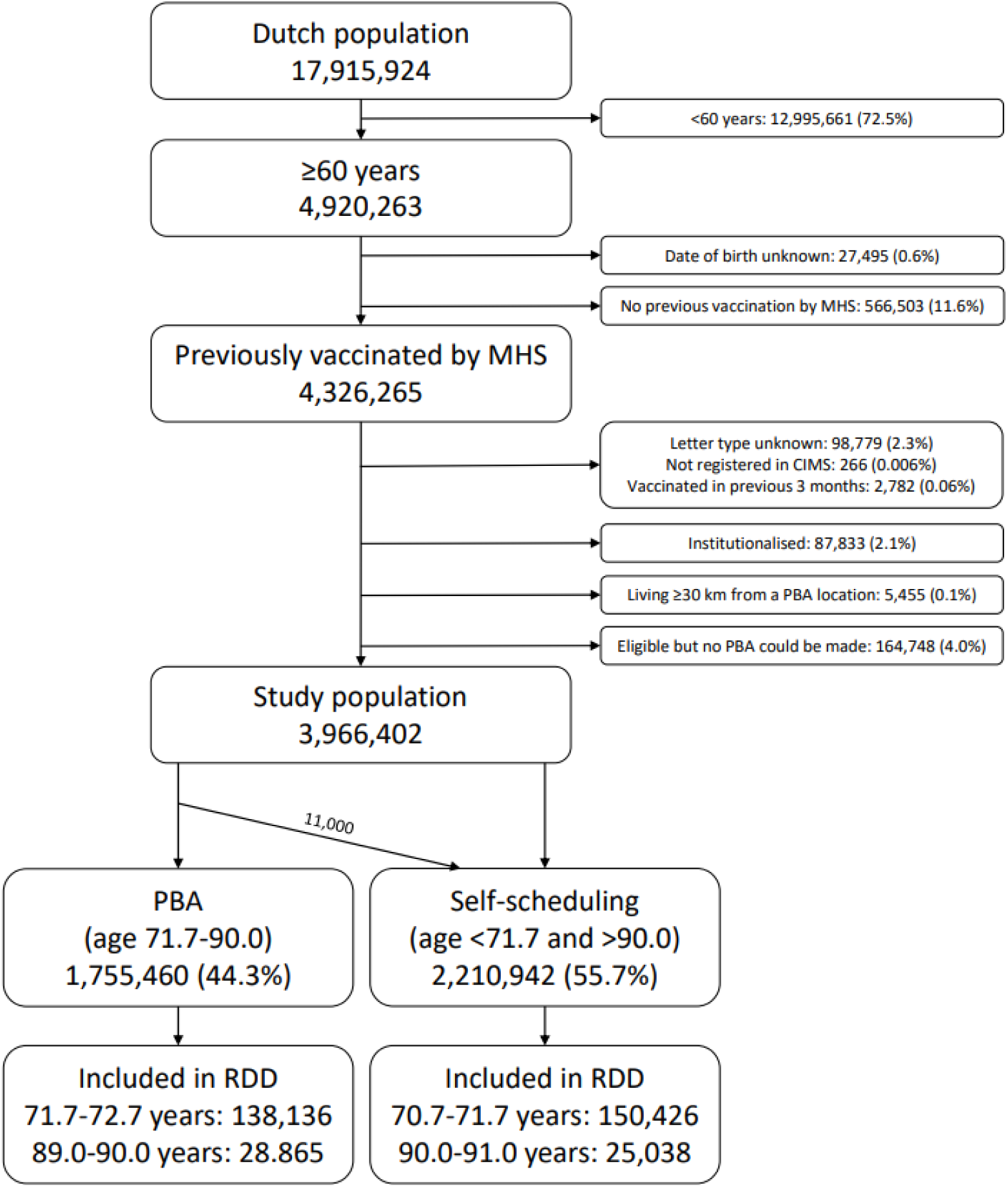
Flow diagram of study population selection *CIMS: COVID-vaccination Information and Monitoring System, MHS: municipal health services, PBA: pre-booked appointment, RDD: regression discontinuity design. Percentages were calculated using the previous population numbers minus the persons excluded in the step(s)/block(s) before. Ages are those on December 31, 2023*.

Therefore, all persons living more than 30 kilometres from the nearest PBA location that was open for at least 10 days were excluded from the analyses to ensure comparability. All institutionalised persons were excluded from the analyses as vaccination was usually organised differently for them. Also, persons with an unknown month of birth were excluded from the analyses. After exclusions, the study population consisted of 3,966,402 persons (Figure 1).

People eligible for PBA who pro-actively booked a vaccination appointment themselves before even receiving an invitation letter, would still receive the PBA letter, even though they were probably already vaccinated at the time they received the letter. This caused confusion in these persons and therefore, in the last batch of PBA letters (i.e., letters for those born between 01-11-1950 and 01-05-1952, scheduled to be sent in week 46), persons who had already booked an appointment themselves were filtered out and instead received a letter to self-schedule an appointment (which they had already done). This applied to 11.000 persons, which was around 5% of persons in this batch. This would disturb the analyses, as these persons intended to get vaccinated, irrespective of the letter type. Also, in the previous batches of PBA letters (sent in week 39-45), this was not done. Therefore, to keep persons invited this last batch of invitation letter comparable to persons invited in the previous batches, these 11.000 had to be recoded to from ‘self-scheduling’ to ‘PBA’, to mimic an intention-to-treat analysis. As we could not identify these persons on an individual level, we randomly redistributed 11.000 vaccinated persons from this last invitation letter batch from ‘self-scheduling’ to ‘PBA’.

### Data and data sources

CIMS, maintained by the RIVM, contains information on administered COVID-19 vaccinations from different health care providers. Also, the type of invitation letter is registered here. Persons are registered in CIMS if they had received a COVID-19 vaccination and gave consent to share their information with the RIVM. During autumn 2023, 98% of all vaccinated persons (both above and below 60 years of age) gave consent to be registered in CIMS [29]. Data were extracted from CIMS on July 2, 2024. From CIMS, information was extracted on type of invitation letter (PBA or self-scheduling), vaccination dates (during the autumn 2023 campaign and before), health care provider that administered the vaccine, and a unique identifier to link the records. If multiple vaccinations were registered in the study period, only the first vaccination per person was included.

Statistics Netherlands (CBS) microdata contains individual level data on all Dutch inhabitants. Information was extracted on month and year of birth, sex, postal code, country of origin, household composition, socioeconomic status, car ownership, and urbanisation level (reference date: October 1, 2023). Data were extracted on February 5, 2025. Individual level sociodemographic data from CBS and the data from CIMS were linked using a random non-identifiable unique number in the secured remote access environment of the CBS.

Information on vaccination locations during the 2023 autumn campaign was received from GGD GHOR Netherlands, the umbrella organisation for the 25 MHS. The dataset contained information on addresses and opening days of vaccination locations, as well as type of location (e.g., open for appointments, open for walk-ins, or both).

### Statistical analyses

An RDD was applied to estimate the local average treatment effect (LATE) at both 71.7 and 90.0 years of age thresholds for PBA eligibility (date of birth 01-05-1952 and 01-01-1934, respectively; age calculated on December 31, 2023). In our study, age was used to assign the type of invitation letter a person would receive. Although age is an important predictor of vaccine uptake [30], it was considered an appropriate assignment variable in the RDD, as it is measured on a continuous scale and cannot be manipulated. Persons immediately above and below the age cutoff were assumed to be interchangeable. We applied a sharp RDD by constructing a binomial generalised linear model with vaccination as the dependent variable, and age (in months) and letter type as the independent variables. The identity link function was used to obtain risk differences. Furthermore, triangular weights were applied, to ensure that more weight was given to values close to the cutoff, while less weight was given to values further away. A bandwidth of 1 year above and below the cutoff was used. The effect of age (i.e. the slope) was assumed to be equal below and above the cutoff. To reduce the impact of model misspecification and potential heteroskedasticity, robust standard errors were computed. LATEs were estimated separately at the age thresholds of 71.7 and 90.0 years.

Analyses were done for the overall population, and stratified by sex, COVID-19 vaccination in the 2022 campaign, country of origin, household composition, socioeconomic status, car ownership, distance to vaccination location, and urbanisation level to check if effects differed between subgroups. These variables were identified in previous studies as important determinants of COVID-19 vaccination [30–32]. If information was missing on any of the variables for stratification, these persons were excluded from the stratified analysis. This only applied to 30 persons missing information on urbanisation (0.0008%) and 49.461 persons missing information on socioeconomic status (1.2%). Interaction terms were added to the model to test if the effect of PBA significantly differed between the subgroups. Sensitivity analyses were performed using different bandwidths. Analyses were done in R version 4.4.0.

## Results

The COVID-19 vaccine uptake in autumn 2023 among the study population of 4.0 million non-institutionalised persons aged ≥60 years with at least one prior COVID-19 vaccination by the MHS was 55.0%, which varied by age (Figure 2) and subgroup (Table 1). Although vaccine uptake was similar across most subgroups, vaccine uptake was much lower among those not vaccinated in the previous year versus those who were (17.7% vs. 74.6%), and among those with specific countries of birth outside of the Netherlands.

**Figure 2.**
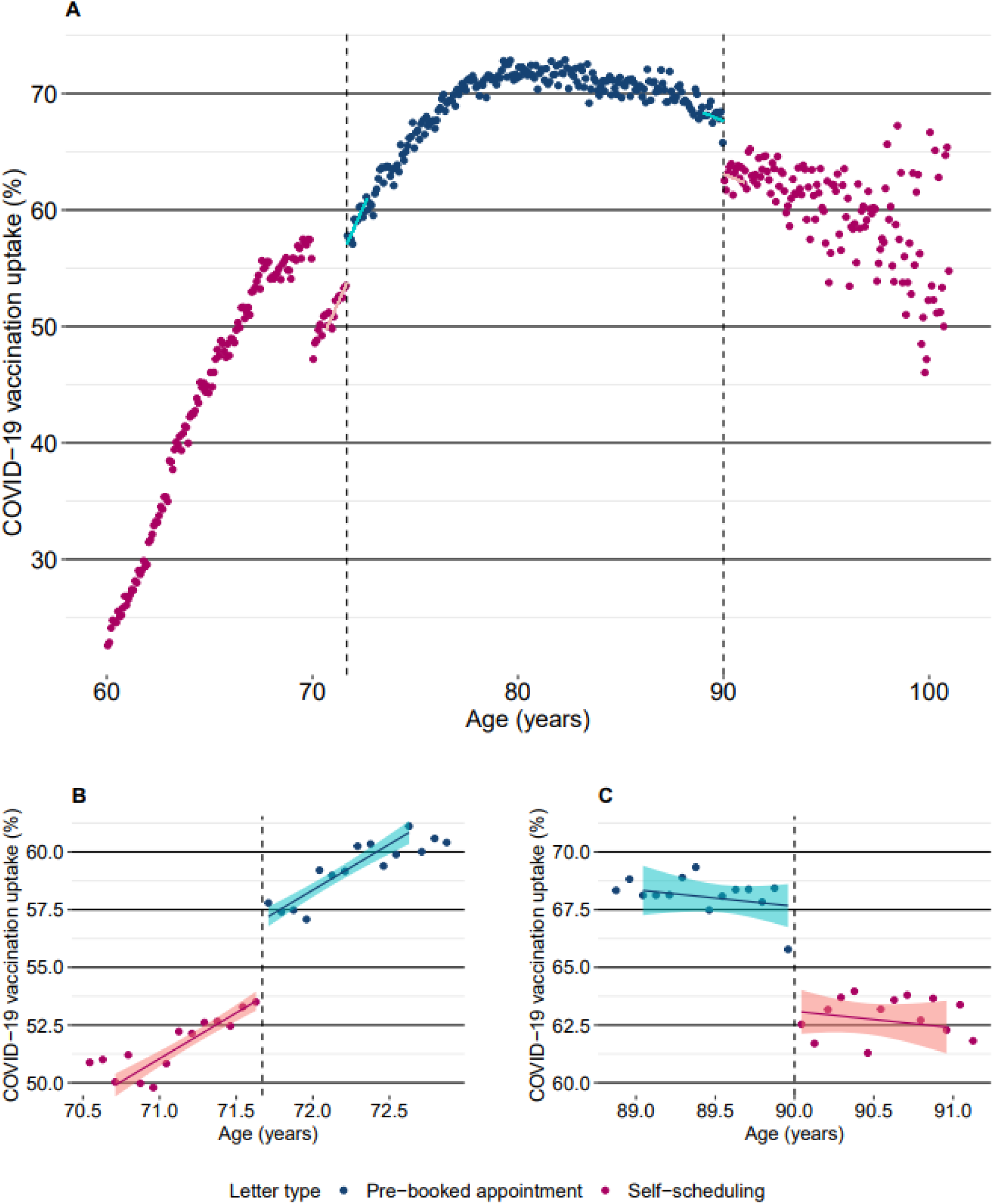
Uptake of COVID-19 vaccination in persons aged ≥ 60 years on December 31, 2023 by letter type with fitted regression lines. Panel A: overall population (N = 3,966,402); Panel B: zoom-in on threshold 71.7 years of age (N = 288,562); Panel C: zoom-in on threshold 90.0 years of age (N = 53,903). *Teal and pink lines: fitted regression lines with 95% confidence interval, using a bandwidth of 1 year before and after the thresholds. Dashed lines: thresholds (age 71.7 and 90.0). NB. An extra discontinuity can be seen at 69 years in panel A. This can likely be explained by the fact that birth cohorts ≥1954 (i.e., 69 years and younger on December 31, 2023) received their invitation up to 2 months earlier than the preceding cohorts; giving them more time to get vaccinated. However, these age groups are not included in our analyses.*

**Table 1.** Vaccine uptake overall and per subgroup of the study population (N = 3,966,402)^a)^

|  |  | ≥60 years |  |  |
| --- | --- | --- | --- | --- |
|  |  | Number | Number vaccinated | Vaccine uptake |
| Overall |  | 3,966,402 | 2,181,267 | 55.0% |
| Sex | Male | 1,900,062 | 1,047,647 | 55.1% |
|  | Female | 2,066,340 | 1,133,620 | 54.9% |
| Vaccinated in 2022 | Yes | 2,599,991 | 1,938,955 | 74.6% |
|  | No | 1,366,411 | 242,312 | 17.7% |
| Country of origin | The Netherlands | 3,379,054 | 1,923,372 | 56.9% |
|  | Child of migrant | 217,003 | 123,177 | 56.8% |
|  | Migrant, Europe | 97,144 | 43,881 | 45.2% |
|  | Migrant, Indonesia | 60,925 | 36,876 | 60.5% |
|  | Migrant, Surinam/Caribbean | 73,918 | 24,036 | 32.5% |
|  | Migrant, Turkey/Morocco | 44,673 | 2,549 | 5.7% |
|  | Migrant, Other | 93,685 | 27,376 | 29.2% |
| Household composition<sup>b)</sup> | Single | 1,213,663 | 627,889 | 51.7% |
|  | Couple | 2,741,177 | 1,547,673 | 56.5% |
|  | Other<sup>c)</sup> | 11,562 | 5,705 | 49.3% |
| Household socio-economic status score<sup>d) e)</sup> | High | 1,335,899 | 707,916 | 53.0% |
|  | Medium | 1,291,017 | 790,217 | 61.2% |
|  | Low | 1,290,025 | 662,613 | 51.4% |
| Household car ownership | Yes | 3,307,740 | 1,860,025 | 56.2% |
|  | No | 658,662 | 321,242 | 48.8% |
| Distance to nearest vaccination location | ≤5 kilometres | 2,250,710 | 1,248,386 | 55.5% |
|  | 5-10 kilometres | 1,249,591 | 688,300 | 55.1% |
|  | >10 kilometres | 466,101 | 244,581 | 52.5% |
| Urbanisation level<sup>f) g)</sup> | Extremely urbanised | 776,370 | 410,475 | 52.9% |
|  | Strongly urbanised | 1,053,565 | 592,183 | 56.2% |
|  | Moderately urbanised | 776,825 | 442,398 | 56.9% |
|  | Hardly urbanised | 673,764 | 379,590 | 56.3% |
|  | Not urbanised | 685,848 | 356,606 | 52.0% |

a) Total group size (i.e., all non-institutionalised persons aged ≥60 years on December 31, 2023 with at least one prior COVID-19 vaccination by the MHS, with no vaccination in the previous 3 months and living ≤ 30 km from a PBA location that was open for at least 10 days). N.B. RDD analyses are based on smaller groups around the thresholds.
b) Children living at home were not taken into account
c) 'Other' includes for example two siblings living together in a household, or a boarder that lives with a family
d) This score represents relative socioeconomic status in comparison with other households, based on wealth, educational level and labour market participation. A higher score indicates more wealthier and higher educated households who have worked for a longer period of time.
e) For 49,461 persons aged ≥60 years (1.2%) the household socio-economic status score was missing, these persons were not included in the analyses
f) Definitions: not urbanised (<500 addresses/km<sup>2</sup>); hardly urbanised (500-999 addresses/km<sup>2</sup>); moderately urbanised (1,000-1,499 addresses/km<sup>2</sup>); strongly urbanised (1,500-2,499 addresses/km<sup>2</sup>); extremely urbanised (≥2,500 addresses/km<sup>2</sup>)
g) For 30 persons aged ≥60 years (0.0008%) urbanisation level was missing, these persons were not included in the analyses

Using a bandwidth of 1 year, a total of 288,562 and 53,903 persons were included in the RDD analyses at 71.7 and 90.0 years, respectively. Overall, the RDD analyses showed a 3.3 (95% CI 2.6-4.1) and 4.5 (95% CI 2.8-6.3) percentage point higher uptake in those with pre-booked appointments compared to self-scheduling at 71.7 and 90.0 years, respectively. Differences were observed between subgroups (Table 2). PBA predominantly led to a higher uptake. The largest effect was observed in children of migrants, at both age cut points (7.1%, 95% CI 3.8 – 10.4; and 11.9%, 95% CI 4.8 – 19.0, respectively). However, in several subgroups no statistically significant effect was seen, while in one (migrants of Turkish or Moroccan origin) a negative effect estimate around the age of 71.7 was observed, with a borderline significantly lower uptake (−3.9%, 95% CI -7.8 – 0.0) of COVID-19 vaccination in the PBA group. This was not observed in this subgroup around the age of 90.0. Adding interaction terms to the models confirmed that statistically significant differences in the effect of PBA existed for different subgroups of country of origin.

**Table 2.** Effect estimates of pre-booked appointments versus self-scheduling on COVID-19 vaccine uptake at two age thresholds overall and per subgroup of the RDD study population

|  |  | 71.7 years (N = 288,562) |  |  | 90.0 years (N = 53,903) |  |  |
| --- | --- | --- | --- | --- | --- | --- | --- |
|  |  | LATE | 95% CI | p-value interaction | LATE | 95% CI | p-value interaction |
| Overall |  | **3.3%** | **2.6 – 4.1** | n/a | **4.5%** | **2.8 – 6.3** | n/a |
| Sex | Male | **4.7%** | **3.5 – 5.8** | **< 0.001** | **4.9%** | **2.1 – 7.7** | 0.36 |
|  | Female | **2.1%** | **1.1 – 3.3** |  | **4.3%** | **2.0 – 6.6** |  |
| Vaccinated in 2022 | Yes | **3.7%** | **2.8 – 4.5** | 0.087 | **5.3%** | **3.5 – 7.1** | 0.78 |
|  | No | **4.1%** | **3.0 – 5.2** |  | **3.6%** | **0.3 – 6.9** |  |
| Country of origin | Born in the Netherlands | **3.9%** | **3.1 – 4.8** | **< 0.001** | **4.4%** | **2.6 – 6.3** | 0.32 |
|  | Child of migrant | **7.1%** | **3.8 – 10.4** |  | **11.9%** | **4.8 – 19.0** |  |
|  | Migrant, Europe | 2.3% | -2.8 – 7.4 |  | 5.8% | -7.9 – 19.6 |  |
|  | Migrant, Indonesia | 5.4% | -0.5 – 11.4 |  | 6.4% | -4.9 – 17.8 |  |
|  | Migrant, Surinam/Caribbean | 2.9% | -2.3 – 8.1 |  | 1.4% | -19.1 – 21.9 |  |
|  | Migrant, Turkey/Morocco | **-3.9%** | **-7.8 – 0.0** |  | 4.4% | -12.2 – 21.0 |  |
|  | Migrant, Other | 2.8% | -2.1 – 7.6 |  | 8.1% | -17.6 – 33.8 |  |
| Household composition<sup>a)</sup> | Single | **5.3%** | **3.7 – 6.8** | 0.091 | **5.5%** | **3.3 – 7.7** | 0.23 |
|  | Couple | **2.6%** | **1.6 – 3.5** |  | 2.0% | -1.0 – 5.0 |  |
|  | Other<sup>b)</sup> | 5.7% | -9.1 – 20.6 |  | 19.8% | -33.6 – 73.2 |  |
| Household socio-economic status score<sup>c) d)</sup> | High | **2.9%** | **1.4 – 4.5** | 0.082 | -2.3% | -8.8 – 4.3 | 0.51 |
|  | Medium | **4.2%** | **3.0 – 5.4** |  | **5.8%** | **2.8 – 8.8** |  |
|  | Low | **2.7%** | **1.3 – 4.0** |  | **4.4%** | **2.1 – 6.7** |  |
| Household car ownership | Yes | **3.2%** | **2.4 – 4.1** | 0.88 | **4.0%** | **1.6 – 6.5** | 0.43 |
|  | No | **4.5%** | **2.4 – 6.6** |  | **4.8%** | **2.3 – 7.3** |  |
| Distance to nearest vaccination location | ≤5 kilometres | **2.0%** | **1.0 – 3.1** | **<0.001** | **5.8%** | **3.5 – 8.2** | 0.29 |
|  | 5-10 kilometres | **5.0%** | **3.6 – 6.3** |  | 2.1% | -1.0 – 5.3 |  |
|  | >10 kilometres | **5.2%** | **3.0 – 7.4** |  | **5.2%** | **0.0 – 10.4** |  |
| Urbanisation level<sup>e) f)</sup> | Extremely urbanised | **3.0%** | **1.2 – 4.8** | **<0.001** | **4.0%** | **0.1 – 8.0** | 0.30 |
|  | Strongly urbanised | **3.3%** | **1.8 – 4.9** |  | **3.3%** | **0.1 – 6.6** |  |
|  | Moderately urbanised | **2.0%** | **0.2 – 3.7** |  | 3.7% | -0.3 – 7.7 |  |
|  | Hardly urbanised | **3.2%** | **1.3 – 5.1** |  | **5.6%** | **1.4 – 9.9** |  |
|  | Not urbanised | **5.9%** | **4.1 – 7.8** |  | **7.8%** | **3.1 – 12.5** |  |

LATE: Local Average Treatment Effect, 95% CI: 95% Confidence Interval, n/a: not applicable
a) Children living at home were not taken into account
b) 'Other' includes for example two siblings living together in a household, or a boarder that lives with a family
c) This score represents relative socioeconomic status in comparison with other households, based on wealth, educational level and labour market participation. A higher score indicates more wealthier and higher educated households who have worked for a longer period of time.
d) For 1.2% of the persons aged ≥60 years the household socio-economic status score was missing, these persons were not included in the analyses.
e) Definitions: not urbanised (<500 addresses/km<sup>2</sup>); hardly urbanised (500-999 addresses/km<sup>2</sup>); moderately urbanised (1,000-1,499 addresses/km<sup>2</sup>); strongly urbanised (1,500-2,499 addresses/km<sup>2</sup>); extremely urbanised (≥2,500 addresses/km<sup>2</sup>)
f) For 0.0008% of the persons aged ≥60 years the urbanisation level was missing, these persons were not included in the analyses
**Bold** indicates statistically significant effect estimates (≠ 0) or interaction p-values

Furthermore, statistically significant differences in the effect of PBA between men and women around the age of 71.7 were observed, with PBA being more effective in men compared to women. Also, significant differences were seen in distance to vaccination location and level of urbanisation, both around the threshold of 71.7 years. The further away the vaccination location was, or the less urbanised the neighbourhood was where the person lived, the bigger the effect of PBA. This trend for urbanisation level was also seen around 90.0 years, although not statistically significant. For all other subgroups and age thresholds no significant differences in the effect of PBA on uptake were seen.

In Figure 3, the uptake over time is shown for the one-year age groups around the thresholds that are included in the RDD analyses. The letter seems to be the most important cue to action, as in the weeks after sending the letters the vaccine uptake rises the most, especially for those with a pre-booked appointment.

**Figure 3.**
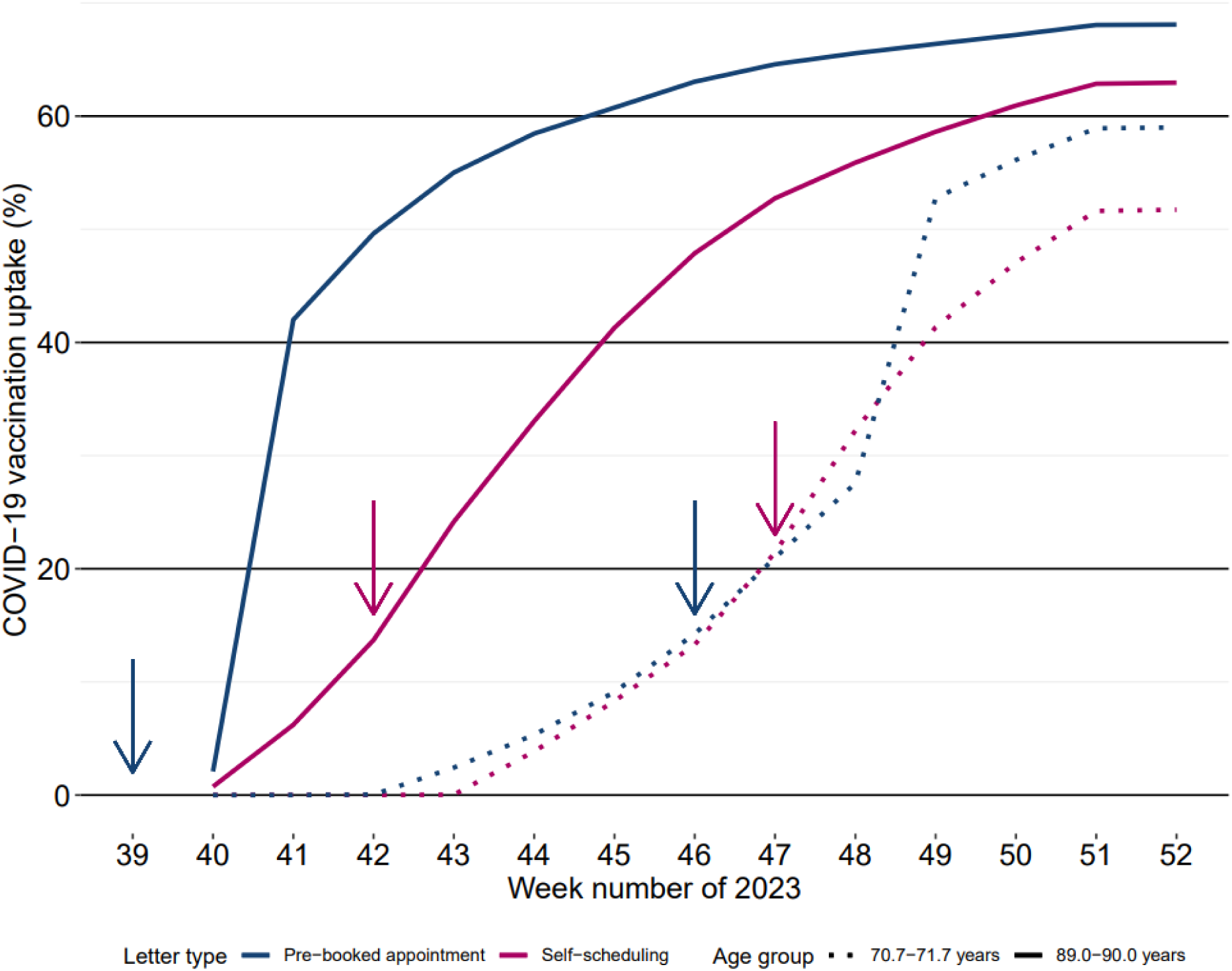
Uptake of COVID-19 vaccination over time in autumn 2023 for one year age groups around the thresholds. *Vaccination started on October 2, 2023 (week 40) and ended on December 31, 2023 (week 52). Arrows: week of sending of letters for the specific age group*.

In Supplementary Table 1 sensitivity analyses are reported, using different bandwidths to estimate the effect of PBA. Changing the bandwidth hardly influenced the results. All positive effect estimates were still present and significance levels were similar. However, effect estimates tended to be slightly lower with broader bandwidths. Although the effect estimates for migrants from Turkey/Morocco was now negative in all sensitivity analyses and at both age cut points, it was not significant anymore when using broader bandwidths. In Supplementary Figure 1 and Supplementary Figure 2 different checks are done on the validity of the RDD assumptions, showing no discontinuity in the other variables.

## Discussion

We found that overall pre-booked appointments were associated with a 3.3% and 4.5% higher uptake of COVID-19 vaccination than self-scheduling at 71.7 and 90.0 years of age, respectively. Since we used an RDD, this can be interpreted as a causal effect [24–27]. Thus pre-booked appointments are thought to be effective in increasing uptake in this context. Our study is the first to evaluate the effect of PBA using a nationwide population registry.

Combining our results with those of Sanders et al. [22], we find that people who have a moderate intention to get vaccinated, but no perception of strong urgency, may particularly be activated to comply with the pre-booked appointment. This might be due to a reduction of the hassle of planning, looking ahead and considering whether to go or not. For that group PBA reduces the intention-behaviour gap [12]. Following these combined results, more age groups (i.e., persons aged 60-90 years) received a PBA letter in the COVID-19 vaccination campaigns in autumn 2024 and 2025 in the Netherlands.

Our results are in line with several other studies that applied PBA for COVID-19 vaccination and also found positive results on vaccine uptake [14–17]. Effect sizes ranged from 2 to almost 12 percentage points. However, these studies differ in the phase of the pandemic and vaccination programme, population demographics, and methods used. Also, the mode of communication of the intervention and control (e.g., letter, text message or general communication) varied between the studies.

In our stratified analyses, we noticed that the effect of PBA did not differ much between subgroups, comparable to Varotsis et al. [17] While we found a negative effect around the age of 71.7 in the stratum of migrants from Turkey/Morocco, e.g., PBA leading to a lower uptake, this should be interpreted with caution. The COVID-19 vaccination uptake in earlier vaccination rounds was lower among migrants [30–32]. As we performed our study among individuals who received at least one earlier vaccination, the effect of PBA refers to a selection of the total migrant population. Also, the estimate was borderline significant in our main analysis; it was no longer significant in the sensitivity analyses. Furthermore, it should be noted that persons with a missing date of birth were excluded. Although overall only a small fraction (0.6%) had a missing date of birth, this mainly concerned migrants (>90%). If a negative effect would exist, this poses challenges in terms of equity of the vaccination programme [33]. If PBA leads to an even lower uptake in certain subgroups with already a relatively low uptake (such as migrant populations), disparities likely become even larger. This echoes previous evidence that interventions aimed at changing a default may not be equally effective across all demographic groups, particularly where baseline intention is low [20, 34]. As such, it indicates that PBA should be implemented with attention to equity, and, where possible, targeted to specific (sub)populations. The underlying reasons for this possible reactance were, however, not studied and should be further investigated to serve this population better.

Adding interaction terms to the model revealed that the effect estimate was significantly different in men compared to women, with a higher effect of PBA in men at the age of 71.7 years. Furthermore, pre-booked appointments were more effective in persons living further away from a vaccination location and persons living in areas that were not urbanized, although the former was only seen at 71.7 years. This suggests that facilitating the process by pre-booked appointments increases the uptake of specific groups, despite the barrier of distance or travel time. As expected, there is a large overlap between these categories. For example, people living in more urbanised areas generally had a shorter distance to a vaccination location, and vice versa (data not shown). Sensitivity analyses using different bandwidths hardly affected the effect estimates, thus showing robustness of our results. Overall, no significant interaction effects were found around the age of 90.0. This could be due to the smaller population size at this age.

In our study we used observational data. The benefit of these data is that the results better reflect effect sizes as they might occur in daily practice. However, due to this, several limitations arose when analysing the data. Choices made in the course of the vaccination campaign were mostly influenced by logistical and practical considerations, not scientific accuracy. For example, 11.000 persons who already booked an appointment themselves were filtered out of the last batch of PBA letters, and received an invitation to self-schedule an appointment. Unfortunately, it was no longer traceable on an individual level who this applied to. Therefore, we randomly redistributed 11.000 vaccinated persons from self-scheduling to PBA. Although this will give a better reflection of the overall effect estimate, a dilution of the subgroup effect estimates around the age of 71.7 years might arise due to the fact that it was unknown which characteristics these 11.000 persons had.

Also, it was decided to only send PBA letters to persons who were vaccinated at least once by the MHS. Furthermore, due to the nature of our design (RDD), results only apply to the age thresholds that were studied. This limits the external validity of our study, as the results cannot be directly extrapolated to non-vaccinated persons or persons of other ages.

Also, it should be noted that the timing of sending the letters might have impacted our results. Letters for those aged 89 years were sent approximately 3 weeks earlier than letters for those aged 90 years. Thus, around the threshold of 90 years, PBA letters were sent earlier than letters with an invite to self-schedule an appointment. This might have led to an overestimation of the effect size at 90 years, as those invited to self-schedule an appointment had less time to do so. Around the age of 71.7 the difference was only one week (with PBA letters being sent earlier), having less of an impact. Also, persons who received their invitation later, might have had a COVID-infection in the meantime. During autumn 2023, COVID-19 circulation started to rise around week 45 (second week of November), coinciding with the letter sending for the 70-73 year old persons [35]. Another limitation is that people who did not give consent to be registered in CIMS during the 2023 autumn campaign were regarded unvaccinated. However, the fraction of people not giving consent was low (<2% for all vaccinations in autumn 2023), thus only having a minor impact on the results. Furthermore, we have no reason to believe that the percentage consent was different in persons with a PBA compared to those who self-scheduled an appointment. Most likely there was no impact on our effect estimates.

In conclusion, from a public health perspective pre-booked appointments were effective, as PBA led to a higher uptake of COVID-19 vaccination than self-scheduling at both age cut points. However, differences between subgroups should be taken into account to increase equity of the vaccination programme. Possible negative responses amongst specific subgroups in the population should be monitored, and taken into consideration in the decision who should be offered PBA, or in the development of targeted letters for specific subgroups.

## Statements

### Ethical statement

This study received medical ethical clearance (EPI-649) by the Centre for Clinical Expertise (KEC) at the RIVM. The KEC is of the opinion that the research does not fulfil the conditions as stated in article one of the Dutch law for Medical Research Involving Human Subjects (WMO) nor in the EU Clinical Trial Directive (2001/20/EC).

### Use of artificial intelligence tools

None declared.

### Data availability

Results are based on calculations by the RIVM using non-public microdata from Statistics Netherlands. Under certain conditions, these microdata are accessible for statistical and scientific research. For further information: https://www.cbs.nl/en-gb/our-services/customised-services-microdata/microdata-conducting-your-own-research

## Acknowledgements

The authors would like to thank Rob van Werven, Ben Bom and William Schuch (RIVM) for their help with the data and analyses, and GGD GHOR Nederland for providing information on vaccination locations.

## Authors contributions

MRH, MSL, JGS and SJMH conceptualised the study. MRH, JvdK, SvdH, HEdM and SJMH designed the study. MRH collected the data. MRH and JvdK did the statistical analyses. MRH wrote the first draft of the manuscript. MRH, JvdK, SvdH, JGS, MSL, MdB, HEdM and SJMH contributed to writing the manuscript. MRH, JvdK, SvdH, JGS, MSL, MdB, HEdM and SJMH approved the final version.

