## Supplementary Material for "The effect of pre-booked appointments on COVID-19 vaccine uptake during the 2023 autumn campaign in the Netherlands: a regression discontinuity analysis"

**Supplementary Table 1. Effect estimates of pre-booked appointments versus self-scheduling on COVID-19 vaccine uptake at two age thresholds and vaccination coverage per subgroup for different bandwidths**

| | | Bandwidth 1.67 years | | | | Bandwidth 2 years[a)] | | Bandwidth 3 years[a)] | |
|---|---|---|---|---|---|---|---|---|---|
| | | 71.7 years (N = 477,822) | | 90.0 years (N = 90,855) | | 90.0 years (N = 109,329) | | 90.0 years (N = 166,223) | |
| | | LATE | 95% CI | LATE | 95% CI | LATE | 95% CI | LATE | 95% CI |
| Overall | | **3.9%** | **3.3 – 4.5** | **4.7%** | **3.3 – 6.0** | **4.7%** | **3.4 – 5.9** | **4.4%** | **3.4 – 5.4** |
| Sex | Male | **5.2%** | **4.3 – 6.1** | **5.1%** | **3.0 – 7.2** | **5.0%** | **3.1 – 7.0** | **4.8%** | **3.3 – 6.4** |
| | Female | **2.7%** | **1.9 – 3.6** | **4.4%** | **2.6 – 6.1** | **4.4%** | **2.8 – 6.0** | **4.1%** | **2.8 – 5.4** |
| Vaccinated in 2022 | Yes | **3.9%** | **3.2 – 4.6** | **4.6%** | **3.3 – 6.0** | **4.6%** | **3.3 – 5.8** | **4.2%** | **3.2 – 5.2** |
| | No | **4.4%** | **3.6 – 5.3** | **4.4%** | **1.8 – 7.0** | **4.5%** | **2.2 – 6.9** | **4.3%** | **2.3 – 6.2** |
| Country of origin | Born in the Netherlands | **4.2%** | **3.5 – 4.8** | **4.5%** | **3.1 – 5.9** | **4.5%** | **3.2 – 5.8** | **4.3%** | **3.2 – 5.4** |
| | Child of migrant | **6.2%** | **3.6 – 8.8** | **10.4%** | **4.9 – 15.9** | **9.8%** | **4.7 – 14.8** | **7.6%** | **3.5 – 11.7** |
| | Migrant, Europe | **4.7%** | **0.7 – 8.7** | 4.7% | -5.9 – 15.3 | 6.3% | -3.4 – 15.9 | 7.6% | -0.1 – 15.4 |
| | Migrant, Indonesia | **6.7%** | **2.1 – 11.2** | 6.2% | -2.4 – 14.8 | 5.5% | -2.3 – 13.4 | 3.5% | -2.8 – 9.8 |
| | Migrant, Surinam/ Caribbean | 3.1% | -0.9 – 7.1 | 1.3% | -14.8 – 17.5 | 1.8% | -13.1 – 16.7 | 4.2% | -8.1 – 16.4 |
| | Migrant, Turkey/ Morocco | -1.5% | -4.4 – 1.3 | -0.4% | -12.8 – 12.0 | -1.5% | -13.2 – 10.2 | -6.8% | -16.9 – 3.4 |
| | Migrant, Other | **4.0%** | **0.2 – 7.8** | 14.8% | -4.3 – 33.8 | 15.6% | -1.7 – 33.0 | 12.5% | -1.5 – 26.6 |
| Household composition[b)] | Single | **5.1%** | **3.9 – 6.2** | **5.1%** | **3.4 – 6.7** | **5.0%** | **3.4 – 6.5** | **4.6%** | **3.4 – 5.9** |
| | Couple | **3.4%** | **2.7 – 4.1** | **3.3%** | **1.1 – 5.6** | **3.6%** | **1.5 – 5.7** | **3.6%** | **1.9 – 5.3** |
| | Other[c)] | 9.8% | -1.7 – 21.2 | 20.8% | -15.8 – 57.4 | 22.5% | -9.7 – 54.8 | 21.9% | -3.8 – 47.6 |
| Household socio-economic status score[d) e)] | High | **3.4%** | **2.2 – 4.6** | 1.6% | -3.3 – 6.5 | 1.8% | -2.6 – 6.3 | 3.2% | -0.3 – 6.8 |
| | Medium | **4.6%** | **3.7 – 5.5** | **6.0%** | **3.7 – 8.3** | **5.8%** | **3.7 – 7.9** | **4.8%** | **3.1 – 6.5** |
| | Low | **3.4%** | **2.3 – 4.4** | **4.1%** | **2.3 – 5.9** | **4.2%** | **2.6 – 5.8** | **4.1%** | **2.8 – 5.4** |
| Household car ownership | Yes | **3.8%** | **3.2 – 4.5** | **4.5%** | **2.7 – 6.4** | **4.5%** | **2.9 – 6.2** | **4.5%** | **3.2 – 5.9** |
| | No | **4.7%** | **3.1 – 6.3** | **4.5%** | **2.6 – 6.5** | **4.5%** | **2.7 – 6.3** | **4.1%** | **2.6 – 5.5** |
| Distance to nearest vaccination location | ≤5 kilometres | **3.0%** | **2.1 – 3.8** | **5.8%** | **4.0 – 7.6** | **5.7%** | **4.1 – 7.4** | **5.2%** | **3.9 – 6.6** |
| | 5-10 kilometres | **5.2%** | **4.1 – 6.3** | **2.7%** | **0.3 – 5.2** | **3.0%** | **0.8 – 5.2** | **3.4%** | **1.6 – 5.2** |
| | >10 kilometres | **4.9%** | **3.2 – 6.6** | **4.7%** | **0.7 – 8.7** | **4.2%** | **0.6 – 7.9** | **3.3%** | **0.4 – 6.2** |
| Urbanisation level[f) g)] | Extremely urbanised | **3.7%** | **2.3 – 5.1** | **4.1%** | **1.1 – 7.2** | **4.1%** | **1.3 – 6.9** | **3.5%** | **1.3 – 5.8** |
| | Strongly urbanised | **4.1%** | **2.9 – 5.3** | **3.7%** | **1.2 – 6.2** | **3.8%** | **1.5 – 6.1** | **4.0%** | **2.2 – 5.9** |
| | Moderately urbanised | **2.6%** | **1.3 – 4.0** | **4.2%** | **1.1 – 7.3** | **4.3%** | **1.5 – 7.1** | **4.4%** | **2.1 – 6.6** |
| | Hardly urbanised | **3.6%** | **2.1 – 5.1** | **4.9%** | **1.6 – 8.2** | **4.6%** | **1.6 – 7.6** | **4.1%** | **1.6 – 6.5** |
| | Not urbanised | **6.1%** | **4.6 – 7.5** | **7.9%** | **4.3 – 11.5** | **7.9%** | **4.6 – 11.2** | **7.0%** | **4.3 – 9.7** |

*LATE: Local Average Treatment Effect. 95% CI: 95% Confidence Interval*

*a) For bandwidths 2 and 3 years, analyses were only done for the age threshold of 90.0 years, as the discontinuity at 69 years (see Figure 2) would bias the results for the age threshold of 71.7 years*

*b) Children living at home were not taken into account*

*c) 'Other' includes for example two siblings living together in a household, or a boarder that lives with a family*

*d) This score represents relative socioeconomic status in comparison with other households, based on wealth, educational level and labour market participation. A higher score indicates more wealthier and higher educated households who have worked for a longer period of time.*

*e) For 1.2% of the persons aged ≥60 years the household socio-economic status score was missing, these persons were not included in the analyses.*

*f) Definitions: not urbanised (<500 addresses/km2); hardly urbanised (500-999 addresses/km2); moderately urbanised (1,000-1,499 addresses/km2); strongly urbanised (1,500-2,499 addresses/km2); extremely urbanised (≥2,500 addresses/km2)*

*g) For 0.0008% of the persons aged ≥60 years the urbanisation level was missing, these persons were not included in the analyses*

***Bold** indicates statistically significant effect estimates (≠ 0) or interaction p-values*

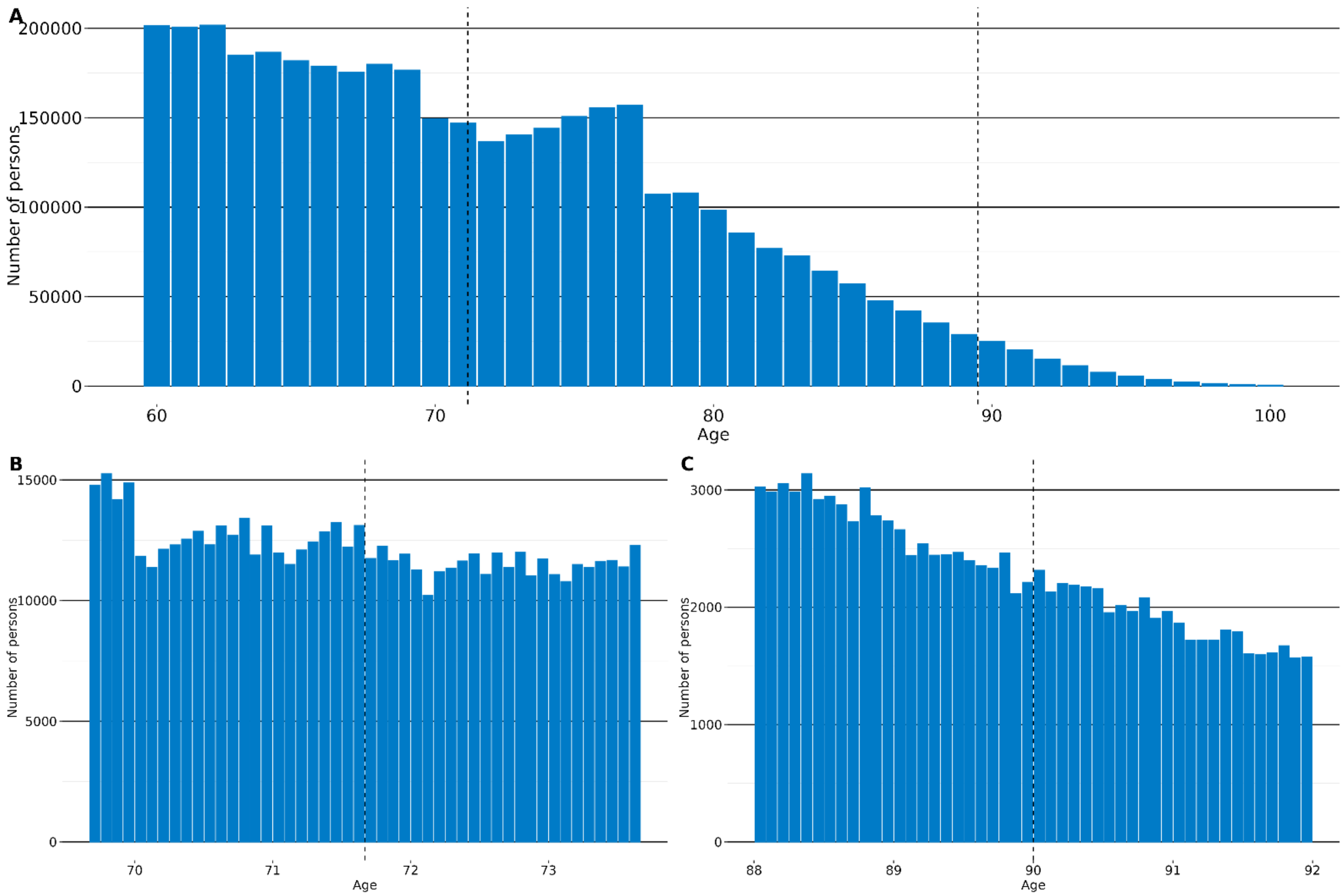


**Supplementary Figure 1. Age distribution of study population**

*A) Complete study population (bin width 1 year), B) Zoom in on threshold 71.7 (bin width 1 month), C) Zoom in on threshold 90.0 (bin width 1 month). Dashed lines: PBA age thresholds. No accumulation of persons at the thresholds can be seen. Discontinuities around 69-70 and 77-78 years of age are known discontinuities in the Dutch population distribution.*

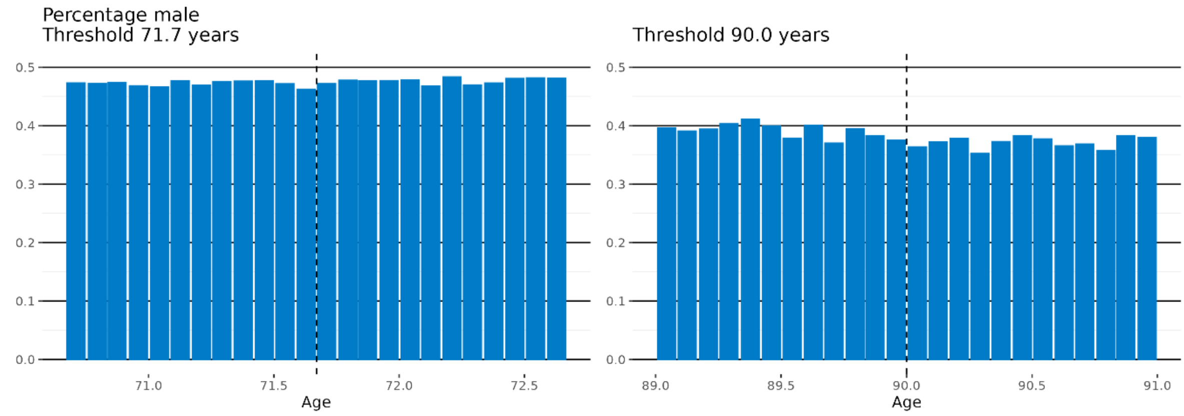
Percentage male
Threshold 71.7 years
Threshold 90.0 years
Age
Age

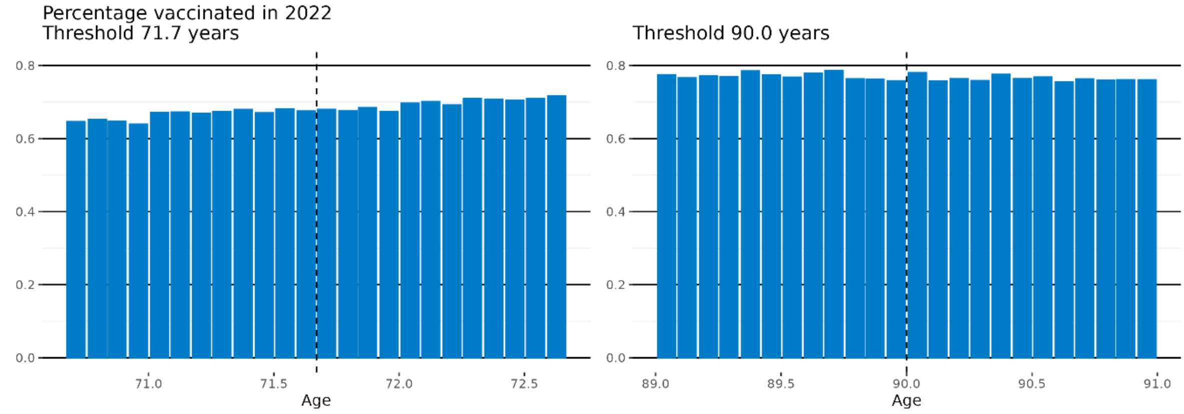
Percentage vaccinated in 2022
Threshold 71.7 years
Threshold 90.0 years
Age
Age

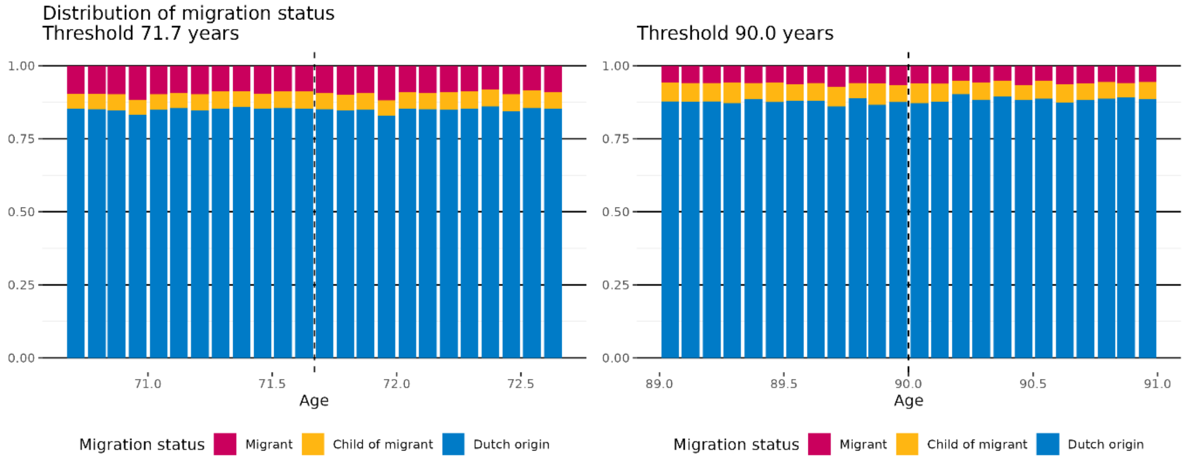
Distribution of migration status
Threshold 71.7 years
Threshold 90.0 years
Age
Age
Migration status
Migrant
Child of migrant
Dutch origin
Migration status
Migrant
Child of migrant
Dutch origin

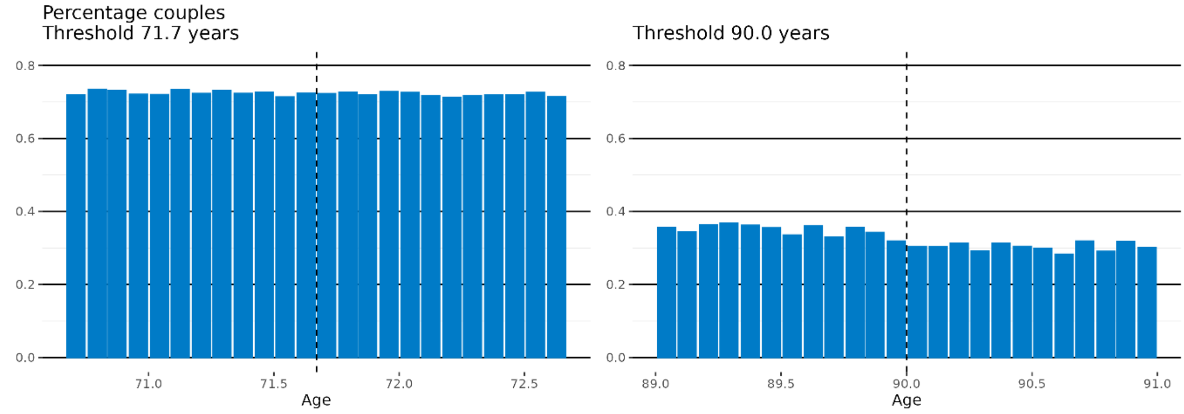
Percentage couples
Threshold 71.7 years
Threshold 90.0 years
Age
Age

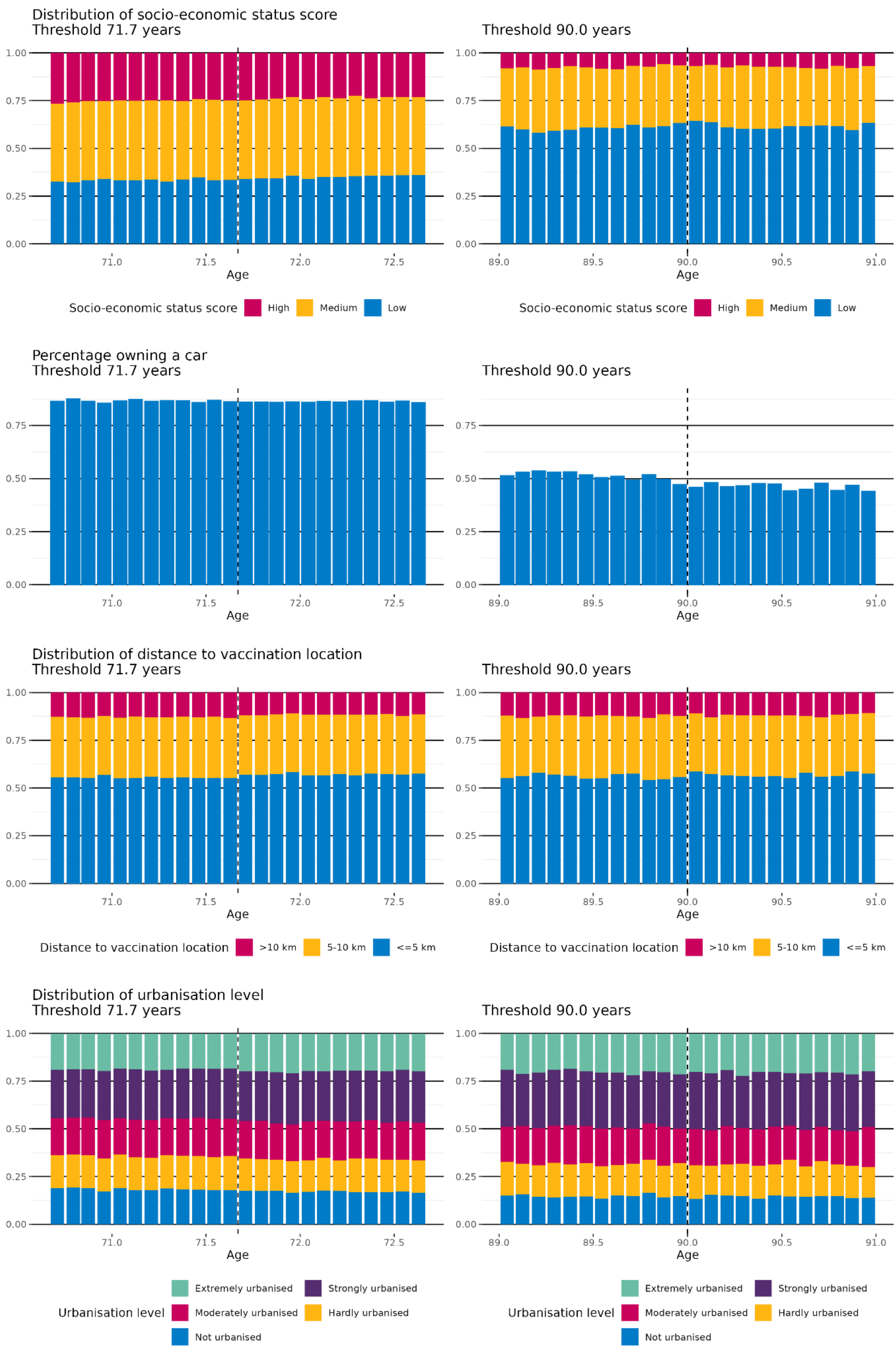

Distribution of socio-economic status score
Threshold 71.7 years
Threshold 90.0 years
Age
Socio-economic status score
High
Medium
Low
Percentage owning a car
Threshold 71.7 years
Threshold 90.0 years
Age
Distribution of distance to vaccination location
Threshold 71.7 years
Threshold 90.0 years
Age
Distance to vaccination location
>10 km
5-10 km
<=5 km
Distribution of urbanisation level
Threshold 71.7 years
Threshold 90.0 years
Age
Urbanisation level
Extremely urbanised
Strongly urbanised
Moderately urbanised
Hardly urbanised
Not urbanised

**Supplementary Figure 2. Distribution of selected baseline characteristics by age**

*No discontinuities of the baseline characteristics can be seen around the RDD thresholds of 71.7 and 90.0 years*
